# 1H NMR metabolomics and lipidomics analysis of neutrophils reveals biomarkers of ageing, inflammageing and frailty

**DOI:** 10.1101/2025.10.29.25339069

**Authors:** Genna Ali Abdullah, Grace Filbertine, Lucy Gill, Asangaedem Akpan, Marie M Phelan, Helen Louise Wright

## Abstract

Neutrophil function declines with age, but little is known about the changes in neutrophil metabolism that take place in older people and people with frailty. We used ^1^H-NMR spectroscopy to investigate the metabolomic and lipidomic profiles of blood neutrophils from healthy younger (HY) and older (HO) people and older people with frailty (FR), to identify metabolites and metabolic pathways altered with healthy ageing and ageing with frailty. We also compared FR metabolites to rheumatoid arthritis (RA) neutrophils to identify markers of inflammation in FR neutrophils. We identified 17 significantly different polar metabolites across the participant groups (ANOVA adj. p<0.05). Three polar metabolites were significantly different in FR vs HO neutrophils, with NADP being higher in FR and lysine and choline being lower. NADP was uniquely elevated in FR compared to HO, HY and RA neutrophils. Seven metabolites were significantly different between HO and HY neutrophils, and only two metabolites were different between FR and RA neutrophils, identifying underlying similarities between FR and RA that may be driven by inflammation. Four lipid peaks were significantly different between FR and HY neutrophils. These were attributed to total cholesterol and omega 6 fatty acids. This is the first study of polar and lipid metabolism in neutrophils from people with frailty, providing evidence that metabolic signalling pathways involved in neutrophil activation, oxidative stress and lipid metabolism are altered during healthy ageing and ageing with frailty, and we identify NADP as a unique biomarker of altered neutrophil function in people with frailty.

## Introduction

Neutrophils are innate immune phagocytic cells that play a key role in protecting the host from infection through the production of reactive oxygen species (ROS), neutrophil extracellular traps (NETs) and proteolytic enzymes such as elastase and collagenase. Neutrophil activation is tightly regulated to prevent damage to host tissues by inappropriate release of cytotoxic molecules, but can become dysregulated in inflammatory diseases such as rheumatoid arthritis (RA) [1]. There is increasing evidence that neutrophils may be activated during inflammation associated with ageing (inflammageing), and this may contribute to the low-grade, chronic inflammatory activation of immune cells and development of cellular senescence and the senescence-associated secretory phenotype (SASP) [2].

It is now demonstrated that, alongside transcription and post-translational modification of proteins, cellular metabolism is a key regulator of immune cell development, differentiation, and activation. For example, resting T-cells switch from oxidative phosphorylation (OXPHOS) to glycolysis and upregulate amino acid consumption upon T-cell receptor (TCR) mediated recognition of an antigen [3]. B-cells skew towards rapid glycolysis once bound to an antigen, producing sufficient ATP to support clonal expansion and large amounts of antibodies [4]. Type 1 (pro-inflammatory) macrophages rely on glycolysis whereas type 2 (anti-inflammatory) macrophages rely on OXPHOS and fatty acid oxidation [5]. Neutrophils demonstrate efficient metabolic flexibility when properly regulated, relying on glycolysis as the fastest pathway to generate ATP. Upon activation, glucose-6-phosphate (G6P), which is an intermediate of the glycolysis pathway, is diverted from the glycolytic pathway into the pentose phosphate pathway to maximise the yield of NADPH which feeds into superoxide production via NADPH oxidase [6]. This is to ensure neutrophils are well-equipped for rapid oxidative burst, bacterial killing and NETs production, eliminating pathogens as quickly as possible.

Metabolites can be reproducibly detected using untargeted nuclear magnetic resonance (NMR) spectroscopy from different components of organismal extracts. Some of these include whole tissues, cell lysates, plasma, serum (different from plasma in that it is cell free and fibrin free), and other bio-fluids such as cerebral spinal fluid, synovial fluid and saliva [7–12]. NMR spectroscopy is mainly used within the field of chemistry and determines metabolites based on their magnetic charge at the atomic level [13]. It can provide information on specific isotopes (typically ^1^H for metabolomics purposes) and aid in distinguishing metabolites by their different molecular structures. ^1^H-NMR provides information based on the ^1^H atoms present in a molecule. NMR is increasingly being used in the life sciences, and is particularly attractive for the use in clinical metabolomics due to the non-destructive and highly-reproducible analysis pipeline [14].

During ageing, metabolic pathways change in immune cells. Parallel to this, metabolite levels also shift. For example, taurine is anti-inflammatory in that it acts as a stimulant of antioxidant pathways [15] and suppresses IL-1β release, restoring ionic homeostasis in immune cells [16]. Blood levels of taurine decline with ageing in humans, mice and monkeys. Taurine deficiency has been proposed as a driver of ageing, with more than 80% difference in circulating taurine levels in elderly individuals compared to younger individuals, and significant differences between sedentary individuals compared to strength trainers, endurance athletes and sprinters [17]. Neutrophils are rich in taurine, and there is an age-related decline of taurine in neutrophils, resulting in weaker antioxidant immune responses [18].

There are currently no studies investigating the metabolite changes that occur within neutrophils from people with frailty. Therefore, the current evidence of changes in neutrophil metabolism in disease is based on reports from different disease states rather than frailty itself. In COVID-19, RA, type 2 diabetes, and cystic fibrosis, neutrophils activation is associated with changes in energy metabolites such as ATP, ADP, and glucose [9], and alterations in metabolites associated with the redox pathway such as glutathione [19] [20]. There are animal studies that give an insight into how neutrophil metabolites may change with age. One study in mice found that there were not only age-related changes but also sex-related metabolic changes in neutrophils which may give insight into why male and females age differently [21]. However, there is no standard scale to score the frailty status of mice and distinguishing them from healthy aged mice, making the study of frailty in mice subjective.

When neutrophils become dysregulated, they cause collateral damage to surrounding tissues which can prolong the inflammatory response unnecessarily, and it is proposed that this can occur in people with frailty [2]. A protocol for extracting polar metabolites from neutrophils and analysis using ^1^H NMR spectroscopy was first reported in 2018 [10], and subsequently applied to neutrophils from inflammatory disease [9]. Although there are a few studies on neutrophil changes with ageing and frailty, there are currently no studies that investigate the changes in metabolites profiles between healthy older people, healthy younger people and older people with frailty. The aim of this study was to perform NMR metabolomic and lipidomic analysis on neutrophils from older people with frailty to identify markers of frailty and to determine the changes in human neutrophils that take place through normal healthy ageing. We also tested our hypothesis that frailty is associated with inflammageing by comparing frail neutrophils with neutrophils from people with RA. Our results indicate that metabolism is altered in human neutrophils with healthy ageing, and with ageing with frailty, with amino acid metabolism being downregulated, and markers of neutrophil activation and oxidative stress becoming elevated. We also show that neutrophils from people with frailty have an inflammatory phenotype, with similarities to neutrophils from people with RA.

## Methods

### Ethics Statement and Study Population

The study was approved by the NRES Southwest Central Bristol Research Ethics Committee for the collection of blood serum from healthy controls and people with frailty (Ref: 21/SW/0093). All participants gave written, informed consent in accordance with the declaration of Helsinki. All participants with frailty fulfilled the Rockwood Clinical Frailty Scale criteria [22] for the diagnosis of frailty (score ≥5). All participants with rheumatoid arthritis fulfilled the American College of Rheumatology 2010 criteria [23]. Participant demographics are shown in Supplementary Table 1.

### Neutrophil isolation

Peripheral blood was collected via venipuncture into lithium-heparin coated vacutainers (Griener Bio-one Ltd). Neutrophils were isolated with Ficoll-Paque as previously described [10]. Erythrocytes were removed by hypotonic lysis with ammonium chloride lysis buffer. Cell counts were adjusted to a final concentration of 5×10^6^cells/mL, and neutrophil purity was assessed by cytospin (routinely >97%).

### Extraction of Neutrophil Metabolites

Both the polar and lipid metabolites were extracted from the same sample from each participant. A concentration of 5×10^6^/mL of freshly isolated neutrophils were pelleted at 1000g for 3 minutes. The supernatant was discarded, and the pellet was flicked to break up the neutrophils. Cells were washed in 1 mL PBS and centrifuged at 20^O^C 1000g for 3 minutes. The PBS supernatant was discarded, and the pellet was placed in a heat block at 100°C for 2 min before being frozen in liquid nitrogen and stored at -80°C. Neutrophil pellets were defrosted on ice, and 500μL of an ice-cold solution of 50% HPLC grade acetonitrile : 50% ice cold double distilled water added. Samples were sonicated with microtip at 20KHz 3 times in an ice bath (2-4 °C)for 30s with 30s rest intervals to prevent sample heating. Samples were vortexed for 1min, then centrifuged at 21,500g for 5 minutes at 4°C. The supernatant (containing polar metabolites) was collected into a new Eppendorf, flash-frozen in liquid nitrogen and lyophilised overnight. The remaining pellet was stored at -80°C for lipid extraction.

### Preparation of polar metabolite NMR sample

Immediately prior to acquisition, 200μL of ice-cold sodium phosphate buffer was added to lyophilised material and vortexed for 1min before centrifugation at 21,500 g for 2 minutes at 4°C. Supernatant (190μL) was transferred into a 3mm NMR tube using a glass Pasteur pipette, and loaded according to predefined, randomised rack positions.

### Preparation of lipophilic metabolite NMR sample

The neutrophil lipid metabolite pellet was lyophilised overnight to remove any remaining supernatant. Then 200µL of 99.9% deuterated chloroform was added to each sample, mixed by pipetting, and transferred into a new Eppendorf tube on ice (4°C). Samples were vortexed for 30s and then centrifuged at 21,500g at 4°C for 5min. Using a glass Pasteur pipette, 190µl of the supernatant was transferred into 3mm NMR tubes leaving any particulate/debris behind.

### NMR Spectral Acquisition Parameters

Samples were analysed on a Bruker Avance IIIHD 700 MHz spectrometer equipped with a 5mm TCI Cryoprobe and a SampleJet automated sample changer, keeping samples chilled (4-10°C) prior to acquisition. The samples were acquired using pulse sequences (vendor supplied cpmgpr1d for the polar samples to attenuate the signals from large molecules and noesygppr1d for lipid samples) enabling quantitative appraisal of small molecule metabolites [24]. Polar metabolite spectra were acquired using 512 scans and lipid spectra were acquired with 256 scans. Polar spectra were acquired at 25°C and lipid spectra were acquired at 15°C.

### Spectral Quality Control (QC)

Spectral QC was performed on each individual spectrum in Topspin 3.5pl6 to appraise water suppression (polar only), baseline, phasing and line width of reference material (TSP (0ppm) or chloroform (7.26ppm) according to best practice [25] with acceptance criteria within one standard deviation of the mean. All failed spectra were re-run up to 3 times within timeframe of 1 week. Spectra were also compared to previously published neutrophil polar spectra [10].

### Metabolite annotation

Polar metabolite annotation was performed using an external library Chenomx (Chenomx Inc., Edmonton, Alberta, Canada) and in-house libraries as described previously [26] (Supplementary Figure 1). The use of an external library such as Chenomx yields a level 2b assignment according to the standard guidelines [25]. In-house libraries of 2D standards (^1^H^1^H COSY spectra) were used to confirm the metabolite annotations leading to an elevated level 1 identification [25]. Lipid reference library was not available to annotate lipid metabolites and therefore, the peaks were numbered, and statistics was run on the numbered unknown peaks. Significant lipid peaks were then matched to published spectral peaks [27, 28]. Spectral integration was performed using tameNMR (www.galaxy.pgb.liv.ac.uk) and representative peaks were selected for each metabolite based on Pearsons correlation on non-overlapped peaks (where possible) for downstream analysis [26].

### Statistical Analysis

For univariate testing, spectra were normalised using probabilistic quotient normalization (PQN) before analysing by one-way ANOVA with application of Benjamini-Hochberg (BH) multiple testing correction and an adjusted *p*-value < 0.05 significance level. Tukey *post-hoc* analysis was performed on between group comparisons. Statistical analyses were performed using R (v4.4.1). For multivariate analysis, normalised spectra were scaled with Pareto scaling before generating partial least squares discriminant analysis (PLS-DA) models (mixOmics package [29]). 2-components PLS-DA models were generated between the experimental groups by dividing the data randomly into 70% training, 30% testing sets and 20 independent models built per comparison. In each independent model performance was assessed by calculating the balanced accuracy, precision, recall and F1-score metrics from the remaining test sample reported as mean and standard deviation. Although PLS-DA performance can be measured via many metrics [30] precision and recall derived from confusion matrices are long established and well reported [31]. Metabolites deemed influential in the model from with variable importance of projection scores (VIP score) >1. PLS-DA model performance was assessed

### Metabolite Pathway Analysis

Over representation analysis and metabolite set enrichment analysis (MSEA) was performed using Metaboanalyst v6.0 [32], against the small molecule pathway database (SMPDB) of 99 metabolite sets based on normal human metabolic pathways. Analysis was filtered for metabolite sets containing at least 3 entries and the pathways were ranked by their FDR-adjusted p-values.

## Results

### Ageing with and without frailty is associated with changes in neutrophil metabolism

^1^H 1D NMR metabolomics analysis of neutrophil metabolites identified 171 polar metabolite peaks, of which 111 peaks were annotated via Chenomx and 60 peaks were attributed to unknown metabolites. 50 representative metabolite peaks were selected based on average Pearson correlation (Supplementary Table 2). Identity key metabolites metabolite peaks (glutathione, lactate, glutamine, and alanine) was confirmed through the use of 2D spectra according to best practice [25].

Application of one-way ANOVA identified 17 significant differences among the 4 participant groups (Table 1). Post hoc analysis revealed 3 significantly different metabolites between people with frailty (FR) and healthy older people (HO), 7 significantly different metabolites between HO and healthy younger people (HY), 15 significant metabolites between FR and HY, and 2 significantly different metabolites between FR and our inflammatory control group of people with rheumatoid arthritis (RA). Boxplots showing normalised abundance of significant metabolites across the four groups are shown in Figure 1A and summarised as a heatmap in Figure 1B. Metabolites that were higher abundance in HY compared to FR included lysine, lactate, leucine, arginine, isoleucine, sarcosine, asparagine, valine, saccharopine, serine, glutathione, and carnosine. In contrast metabolites higher in abundance in FR compared to HY were GTP, NADP, and 3-hydroxyisovalerate. Glutathione and carnosine distinguished HY from all other groups showing a decrease of antioxidant metabolites with age and disease. There were 7 significantly different metabolites between HY and HO, including carnosine, valine, acetamide, glutathione, arginine, serine, and saccharopine which were all higher in HY compared to HO. NADP and isoleucine were the only 2 metabolites significantly different between FR and RA, with NADP higher in FR and isoleucine higher in RA. Lysine, choline and NADP were significantly different between HO and FR. NADP was the only metabolite significantly higher in FR compared to all other groups.

**Figure 1.**
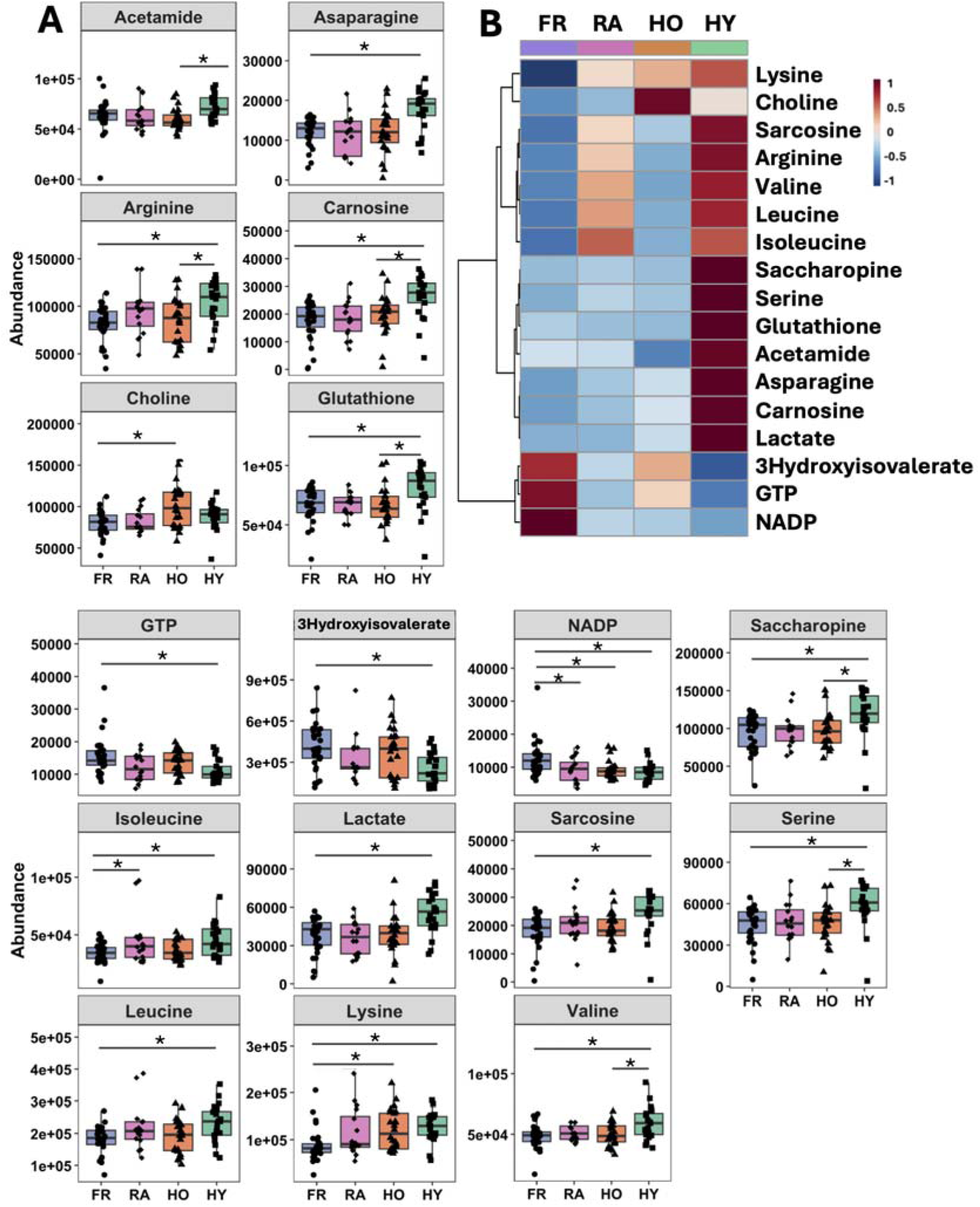
Significantly different metabolites from the 50 CRS-selected representative peaks between at least two groups via one-way ANOVA. (A) Boxplots of significantly different metabolites by ANOVA. Significant differences are indicated with the bars above the boxplots and an asterisk (*) representing the adjusted *p*-value<0.05. (B) Heatmap of significantly different metabolites showing average expression by group, blue=low, red = high. FR = frail, blue circles, n=31; HO = healthy older, orange triangles, n=24; HY = healthy young, green squares, n=21; RA = rheumatoid arthritis, pink diamond, n=16.

**Table 1.**
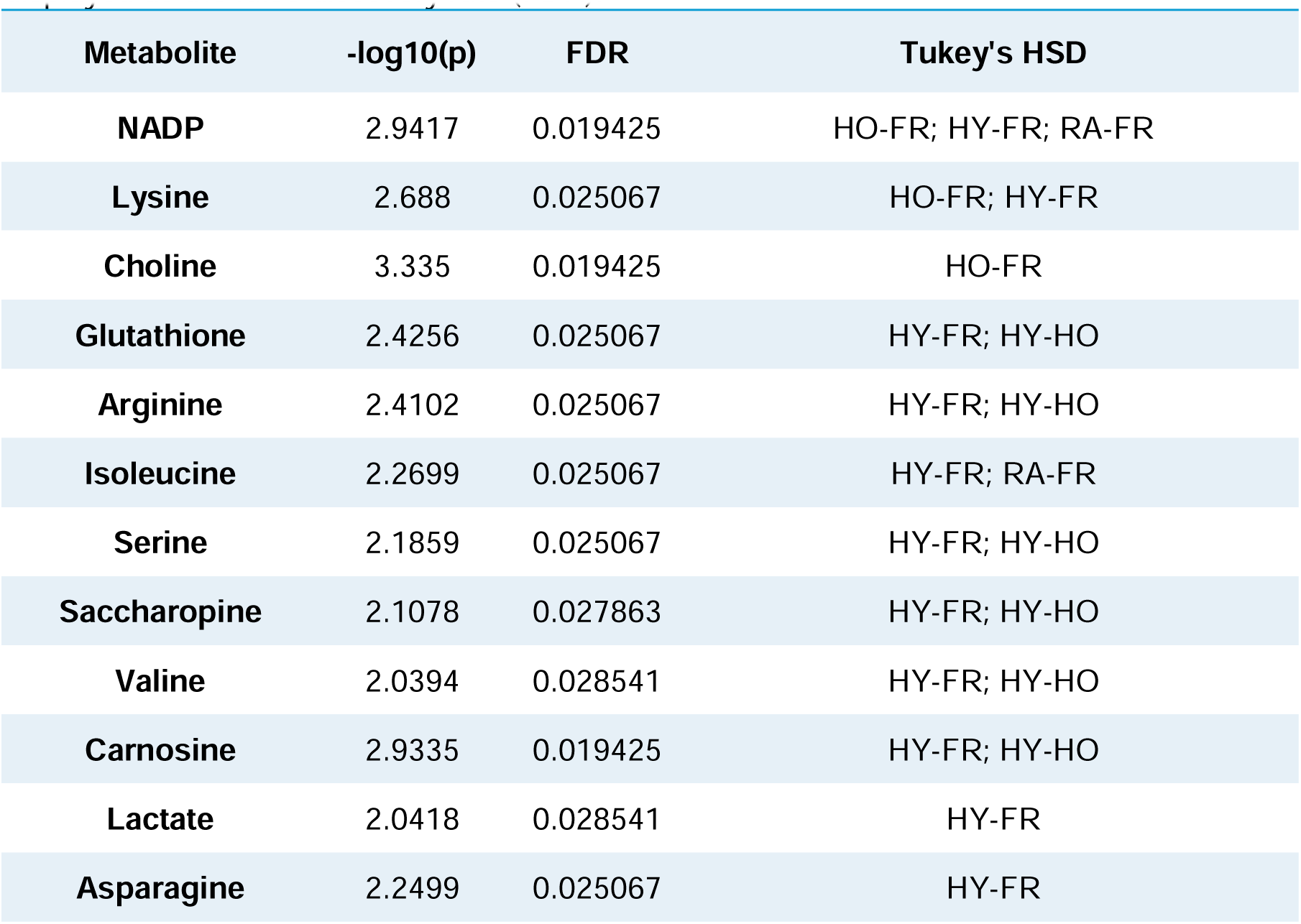
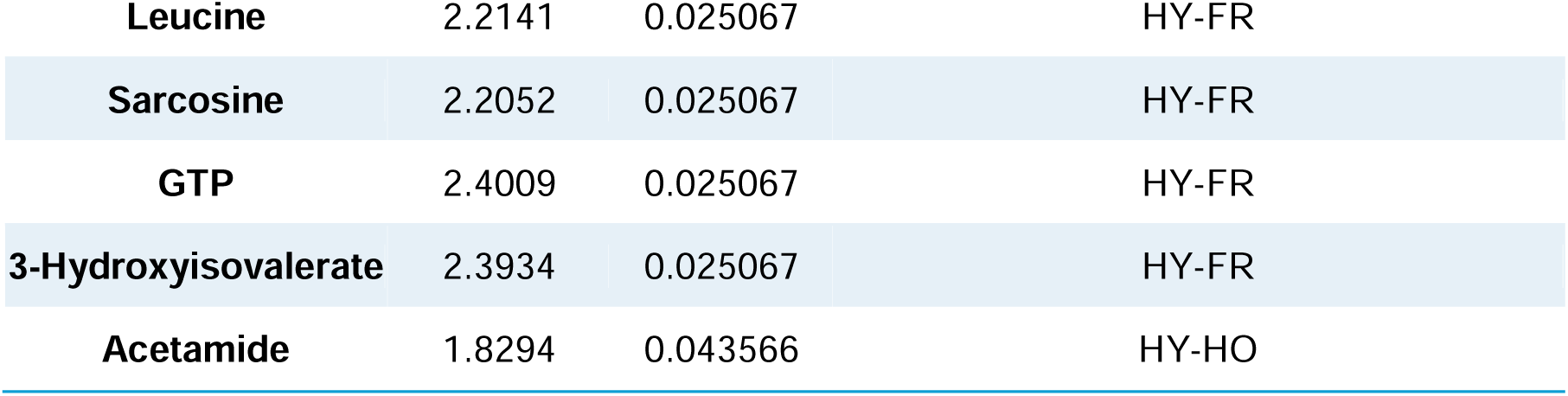
Significant differences between each of the 4 groups analysed by one-way ANOVA with Tukey’s honestly significant difference (HSD) post-hoc tests. The comparisons of interests were FRvsHY, FRvsHO, HOvsHY, and FRvsRA. Adjusted *p*-value displayed as the false discovery rate (FDR).

In order to investigate the difference between FR and the contrasting healthy groups (HO and HY) without any effects from the heterogenous RA group, RA was removed and one-way ANOVA was performed on FR, HO and HY alone. ANOVA identified an additional five metabolites (adj. p<0.05, Supplementary Figure 2). Four of these metabolites, alanine, proline, cysteine, and aspartate were higher in HY compared to both HO and FR. Acetoacetate was higher in both FR and HY compared to HO. We noted that all the amino acids found to be significantly different by ANOVA were increased in the HY group, besides choline and lysine which are increased in HO group compared to FR. This indicates a decline in amino acid abundances in neutrophils associated with ageing.

### Multivariate analysis identifies biomarkers of healthy ageing and ageing with frailty

Supervised multivariate analysis was performed via PLS-DA models to identify important metabolites distinguishing each participant group. A 2-component model of FR vs HO neutrophil metabolites (Figure 2A) had a balanced accuracy of 69.9% (±9.2%) and exhibited moderate precision (63.5% ± 17.1%) and sensitivity (70.2% ± 18.4%). Beyond model performance, the PLS-DA analysis highlighted 17 influential metabolites (VIP>1) discriminating FR and HO which are summarised in a heat map (Figure 2A). Three of these metabolites, NADP, lysine and choline, were also identified as statistically significant (adj.p-value<0.05) through univariate analysis, demonstrating agreement between the two approaches. In order to probe the structure of the data further the VIP>1 metabolites were subset for an unsupervised principal component analysis (PCA) which improved the separation of the groups compared to the entire dataset (Supplementary Figure 3A).

**Figure 2.**
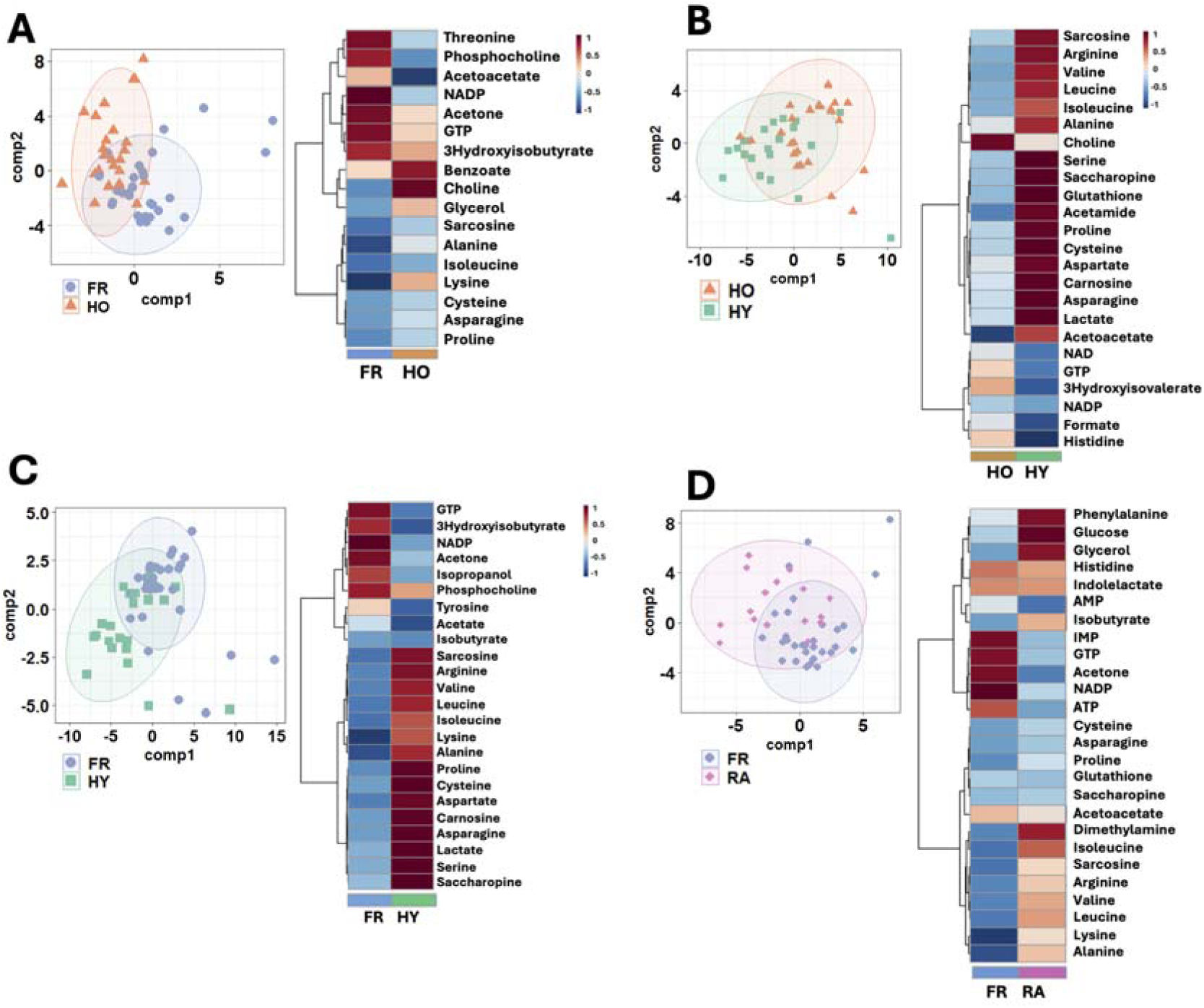
Multivariate analysis of neutrophil metabolites. Two-component PLS-DA of CRS-variable selected metabolites with VIP>1 metabolites summarised in a heatmap for (A) for FR vs HO, (B) HO vs HY, (C) FR vs HY, and (D) FR vs RA. Neutrophils were isolated from participants who were healthy old (HO, orange triangles, n=24), frail (FR, blue circles, n=31), healthy young (HY, green squares, n=21) or rheumatoid arthritis (RA, pink, n=16). For heatmaps, metabolite abundances were log10 transformed and displayed as mean intensities of blue or red. Blue shows lower than mean abundance and red shows higher than mean abundance.

The metabolic changes associated with healthy ageing was also explore with a multivariate 2-component PLS-DA model of HO vs HY (Figure 2B). This had a balanced accuracy of 57.7% (±12.4%) with precision of 52.8% (±20.1%) and sensitivity of 62.3% (±18.1%). These metrics indicated a model with poorer predictive power than between FR and HO. The model identified 23 metabolites with VIP>1, 7 of which were significant by univariate analysis. All 7 metabolites, glutathione, arginine, serine, saccharopine, valine, carnosine and acetamide, were higher in HY compared to HO neutrophils. In order to probe the structure of the data further these metabolites were subset for a PCA and this led to a partial separation of the sample groups (Supplementary Figure 3B).

Multivariate PLS-DA comparing FR with HY neutrophils (Figure 2C), performed better than HOvHY and FRvHO with a balanced accuracy of 72.6% (±10.3%), precision of 67.3% (±17.1%) and sensitivity of 64.7% (±14.3%). This model identified 24 metabolites with VIP>1 (Figure 2C and Supplementary Figure 3C), of which 13 were also significant by univariate analysis. Notably arginine, serine, saccharopine, valine and carnosine were again higher in HY compared to FR neutrophils, suggesting these metabolites may be elevated due to ageing rather than frailty.

We finally modelled the difference between FR neutrophil metabolites and our inflammatory control RA neutrophils using PLS-DA (Figure 2D). This model had a balanced accuracy of 68.5% (±13.3%) with precision of 58.6% (±20.7%) and sensitivity of 66.1% (±23.3%). Twenty-six metabolites exceeded VIP>1 (Figure 2D and Supplementary Figure 3A), of which NADP (higher in FR) and isoleucine (higher in RA) were significant by univariate analysis.

The metabolites consistently reported within via PLS-DA VIP scores provide insight into key metabolites which become altered in neutrophils during healthy ageing and ageing with frailty. Notably, GTP, NADP and acetone were consistently higher in FR compared to HO, HY and RA neutrophils, indicating these metabolites may be potential biomarkers of frailty in human neutrophils.

### Lipid metabolism is altered in frailty

We next performed lipidomics analysis of neutrophils using 1D ^1^H NMR metabolomics. We identified 157 lipid spectral peaks. We did not subset the data for lipidomics analysis as lipid libraries for NMR are not available via publicly available NMR databases (such as Chenomx). Lipid peaks were numbered and grouped based on their characteristics such as lipid polar head group or hydrocarbon tail spectral peaks (Figure 3). Peaks that were significantly different by ANOVA (FDR adjusted P-value <0.05) were further investigated and the lipids were annotated using a previously published study on lipid metabolomics [27]. The lipid and polar metabolites were extracted from the same sample of neutrophils collected from each participant and thus the lipid and polar data are paired.

**Figure 3.**
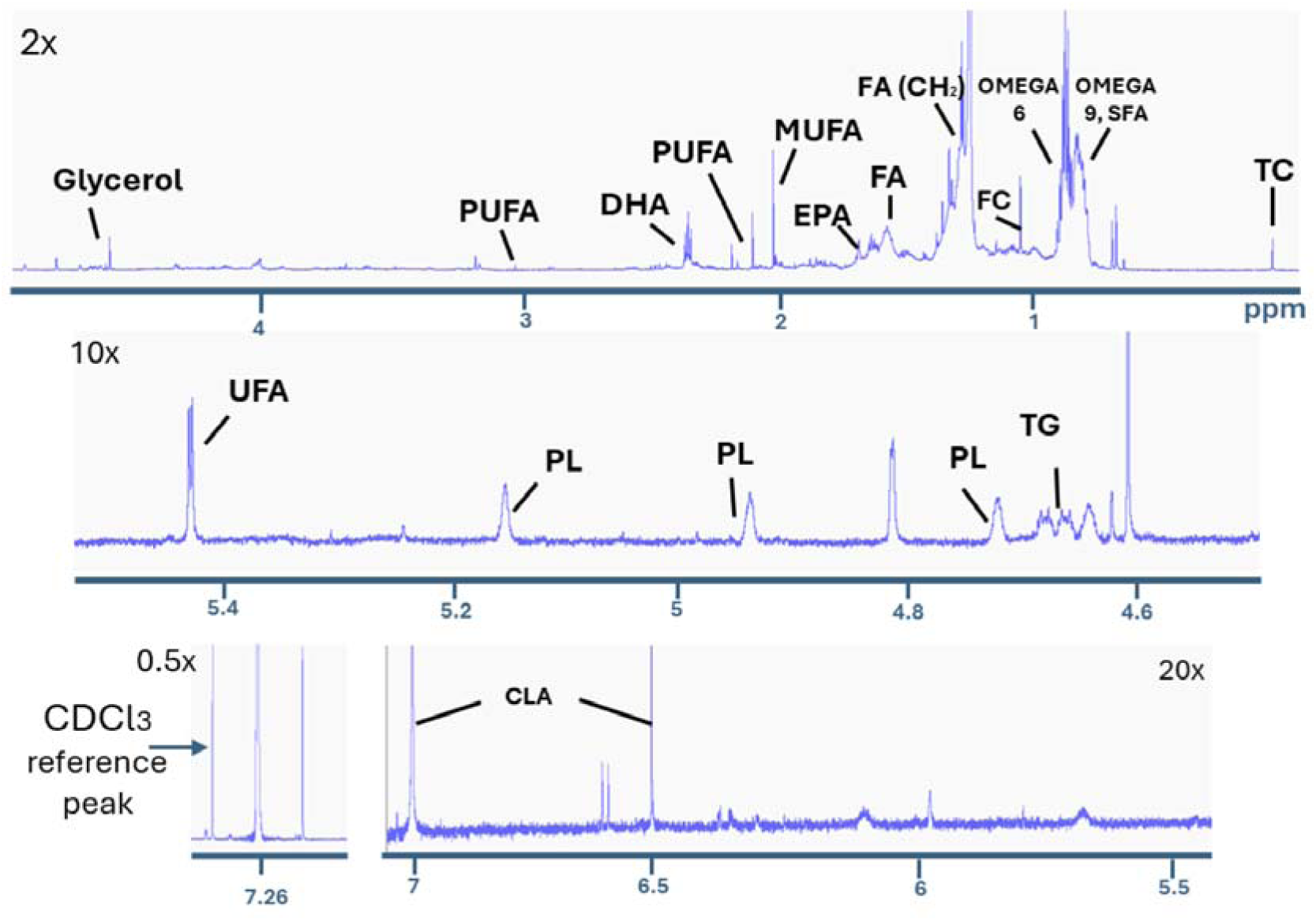
700MHz ^1^H-NMR spectra of lipophilic extracts from human neutrophils. The peaks are labelled as follows: TC, total cholesterol; FC, free cholesterol; TG, triglycerides; FA, fatty acids; SFA, saturated fatty acids; UFA, unsaturated fatty acids; MUFA, monounsaturated fatty acids; PUFA, polyunsaturated fatty acids; EPA, eicosapentaenoic acid; DHA, docosahexaenoic acid; PL, phospholipids; CLA, Conjugated linoleic acid, CDCl3; chloroform, ppm; parts per million [28].

One-way ANOVA on the total number of spectral bins showed there was only one significant bin between FR and HY. A Student’s t-test applied between each of the 4 groups which identified 4 significant lipids bins between FR and HY, and no significant differences between FR and HO, FR and RA, and HY and HO. The significant peaks between FR and HY were tentatively assigned to total cholesterol (boxplot P1) and the omega 6 fatty acid arachidonic acid (AA) (boxplots P75, P77 and P78, Figure 4).

**Figure 4.**
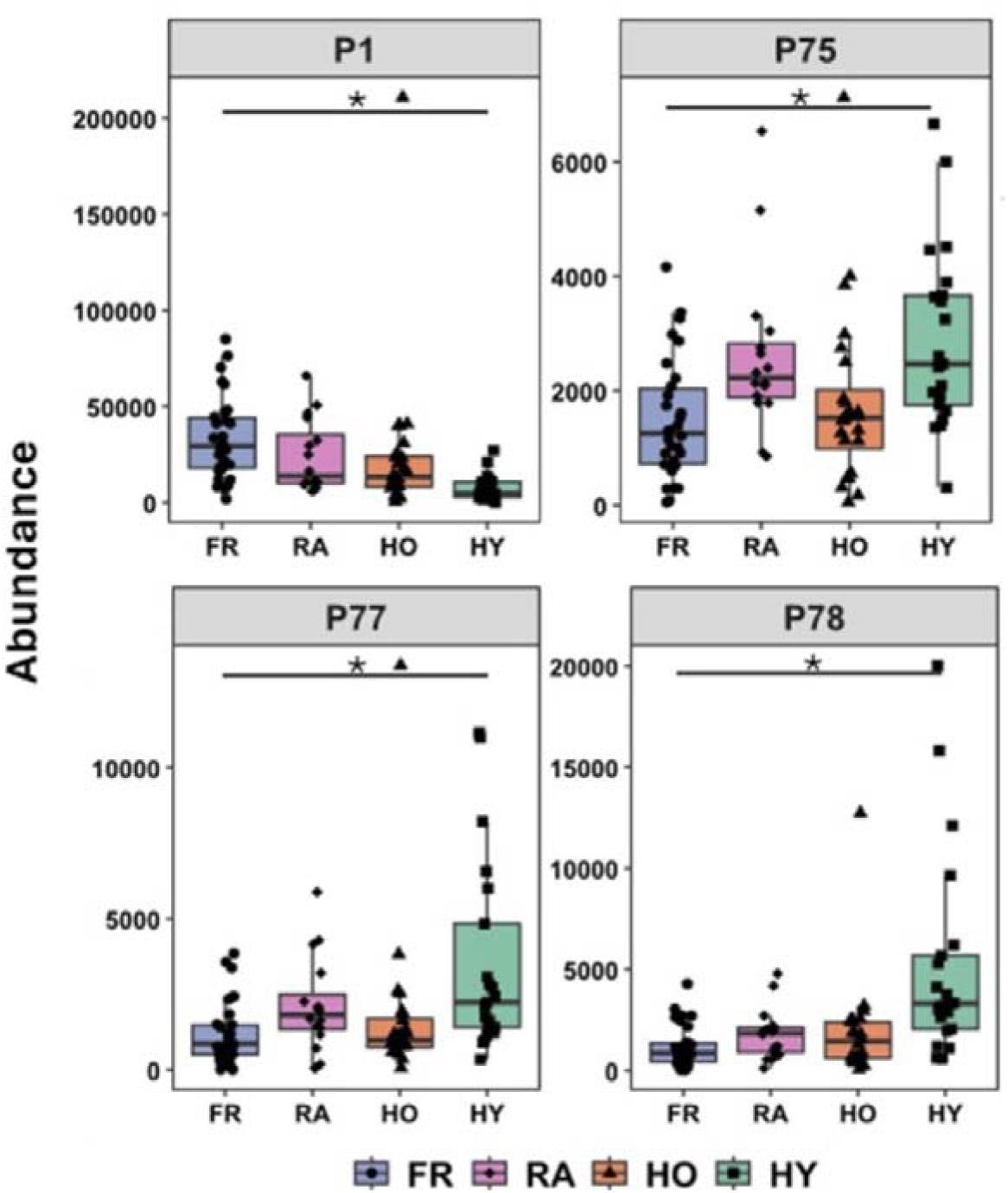
Significantly different lipid metabolite peaks between frail and healthy young neutrophils. Boxplot P1 is assigned as total cholesterol. P75, P77, P78 are omega 6 fatty acids that include arachidonic acid, which is significantly lower in FR compared to HY. FR (blue) and HY (green), p<0.05.

PLS-DA of FR and HO could not be classified above random (balanced accuracy of 47.7% (%11.3%), indicating that the model is not reliable for predicting FR and HO, and that lipid changes for this cohort are mainly associated with age and not frailty status.

In contrast FR and HY had the greatest balanced accuracy of 74.5% (±13.2%, Figure 5A), which correlates with the polar metabolite analyses and also lipid univariate analysis as most significantly different metabolites were between FR and HY. There was moderate classification specifically between the HY and HO (63.7% ±12.1, Figure 5B) balanced accuracy. FR vs RA had a balanced accuracy of 60.4% (±13.3%, Figure 5C). VIPs>1 between FR and HY (Figure 5D) arise from the significantly different peaks (P1, P77, P78) as well as peaks P22-P31 (1.11-1.29ppm) which are attributed to free cholesterol, and these were all higher in HY. We also observed different spectral peaks driving the differences observed between the HO:HY (Figure 5E) and FR:RA groups (Figure 5F) although they are not annotated to specific lipids.

**Figure 5.**
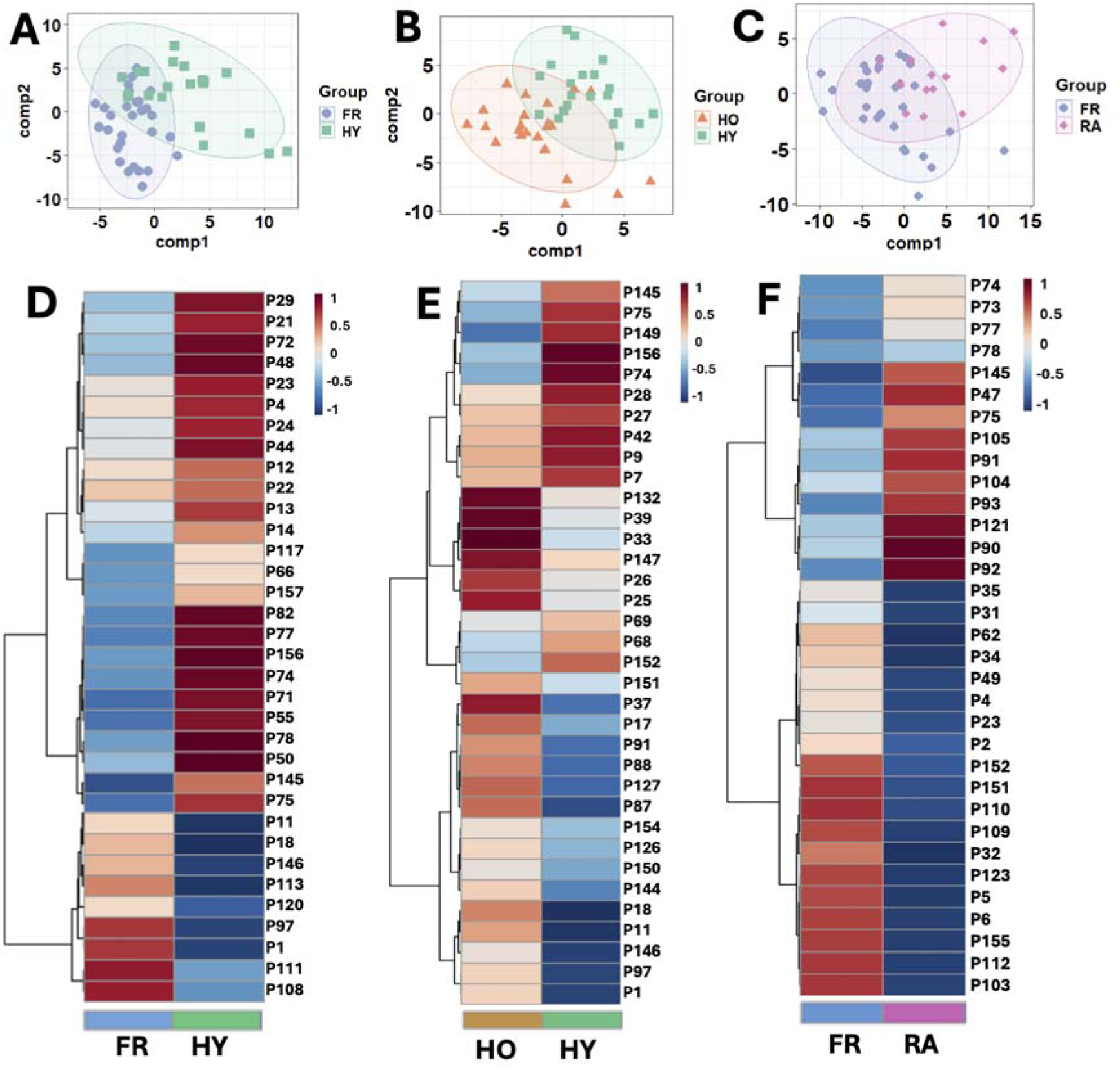
Multivariate analysis of lipid metabolites. PLS-DA scores plots for models with moderate predictive power (A) FR vs HY, (B) HO vs HY, (C) FR vs RA). (B) Top 35 peaks based on the VIP scores (score>1) from the PLS-DA models for neutrophil lipids between each group for (D) FR and HY, (E) HO and HY, and (F) FR and RA. FR and HO was not included as the model could not classify the groups beyond random chance. FR = frail, blue circles, n=31; HO = healthy older, orange triangles, n=24; HY = healthy young, green squares, n=21; RA = rheumatoid arthritis, pink diamond, n=16.

### Pathway Enrichment Analysis reveals differences in neutrophil glutamine and glutamate metabolism in healthy ageing and frailty

Metabolite set enrichment analysis (MSEA) against the small molecule pathway database (SMPDB [33]) of 99 metabolite sets based on normal human metabolic pathways was performed to contextualise the changes in metabolites of interest from this data set. Metabolites of interest were selected from significant differences via ANOVA as well as the metabolites influential in PLS-DA models via VIP score >1. The top 25 pathways enriched by MSEA are shown in Figure 6. Four pathways were significantly enriched between FR and HO which were aspartate metabolism, ketone body metabolism, beta-alanine metabolism and ammonia recycling (Figure 6A, Supplementary Table 3, adjusted p-value < 0.05). Only one pathway, glutathione metabolism was significantly enriched between FR and HY (Figure 6B, Supplementary Table 3, adjusted p-value < 0.05). Only 3 pathways were significantly enriched for HO and HY (Figure 6C, Supplementary Table 3, adjusted p-value < 0.05) which were glycine and serine metabolism, methionine metabolism, and ammonia recycling. The top 10 listed pathways for FR and RA were all significantly enriched (Figure 6D, Supplementary Table 3, adjusted p-value < 0.05) including pyruvate metabolism, valine, leucine and isoleucine metabolism, glutathione metabolism, and thiamine metabolism.

**Figure 6.**
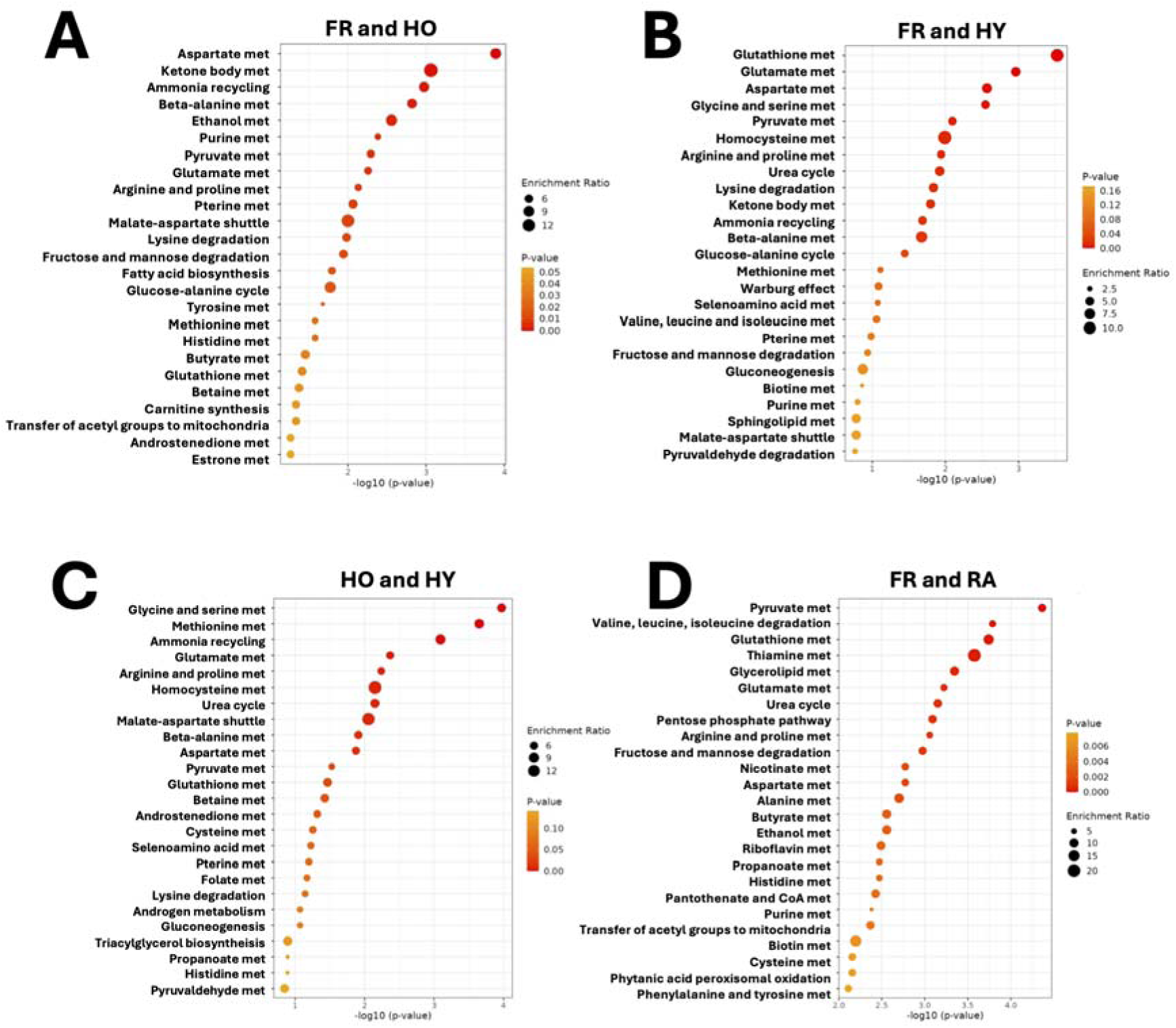
Metabolic pathway enrichment analysis. Metabolite pathway enrichment analysis for the metabolites of interest as determined via VIP score for (A) FR and HO, (B) FR and HY, (C) HO and HY, and (D) FR and RA.

Based on enrichment analysis and neutrophil metabolism a set of pathways are proposed as altered within neutrophils from people with frailty. This includes glutathione metabolism, which was significantly lower in FR neutrophils, and glutamate metabolism which was identified as a significant pathway between FR and HY and HO and HY neutrophils. Whilst glutamate itself was not significantly different between groups, the metabolites saccharopine, arginine and proline were all higher in HY compared to FR and HO and these metabolites feed into the glutamate synthesis pathway (summarised in Figure 7).

**Figure 7.**
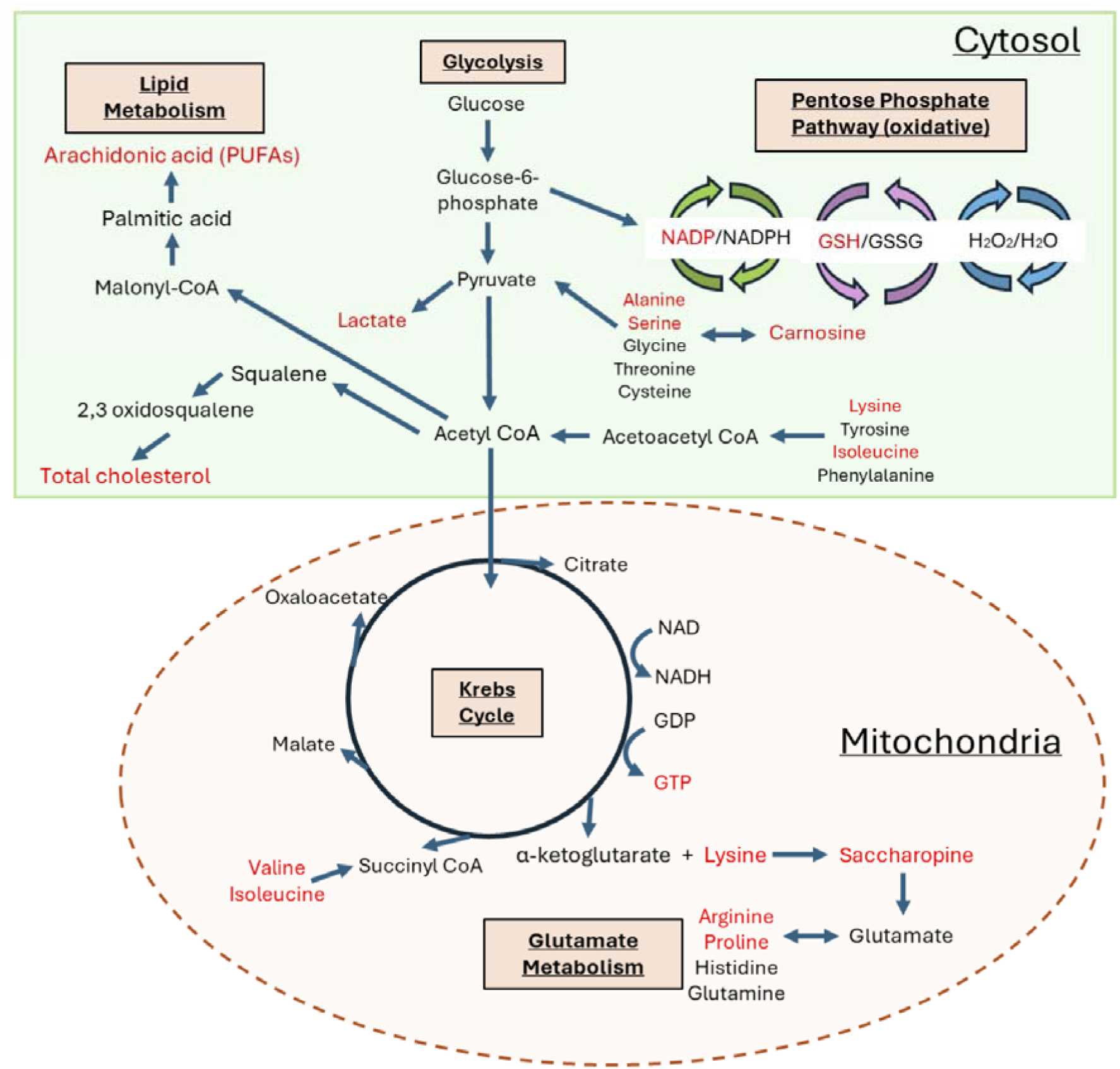
Proposed metabolic pathways that are altered in neutrophils from people with frailty. Neutrophil metabolites altered in the cytosol and mitochondria (highlighted in red). There have been mixed reports on whether neutrophils utilise the Krebs cycle, therefore, these metabolites could have an alternative source that has not yet been defined.

## Discussion

In this study we have used ^1^H NMR metabolomics to study the differences in neutrophil metabolites in people with frailty compared to healthy older and healthy younger healthy controls and people with RA. Both polar and lipid metabolites were analysed in the same samples, providing a novel approach to understanding altered neutrophil metabolism in frailty and ageing. We identified dysregulation in a number of related metabolic pathways, notably ones affecting oxidative stress, immune function and lipid metabolism.

Our analyses of polar neutrophil metabolites identified NADP as the only metabolite statistically significantly higher in FR neutrophils compared to all other groups, and therefore this could represent a novel biomarker of immune frailty. As well being a metabolite of respiration, NADP represents the oxidised form of NADPH, and is involved in pathogenic neutrophil oxidative reactions. The overproduction of NADP and NADPH can promote the production of ROS which, without a pathogenic stimulus, unnecessarily disrupts and damages healthy cells [34], and in neutrophils ROS also fuel production of tissue-damaging NETs. It is notable that in this study, neutrophil NADP in frailty is higher even than in RA neutrophils, an inflammatory disease typically associated with excessive neutrophil reactive oxygen species production [1, 9].

Glutathione and carnosine were significantly higher in HY compared to HO and FR, showing a difference with age rather than frailty status. Both metabolites have antioxidant functions. Glutathione has a well-established protective role, neutralising oxidative stress and aiding in the resolution of inflammation [35]. Glutathione is synthesised from cysteine, glutamate, and glycine. Supplementation of a combined form of glycine and N-acetylcysteine (termed GlycNAC) significantly increased glutathione levels in whole blood of older adults (mean age 65) with high oxidative stress at baseline [36]. A recent randomised, placebo-controlled trial showed that GlycNAC supplementation for 16 weeks increased parameters of physical function such as gait speed, BMI and waist circumference [37]. They also found improvements in several hallmarks of ageing affecting mitochondrial dysfunction, inflammation, genomic damage, and cellular senescence compared to baseline in the participant’s skeletal muscle and urine [37]. A previous study in neutrophils found that glutathione and NAC supplementation reduced the levels of ROS produced when stimulated with LPS to near baseline levels, although the age of the donors was not stated [38]. There are reported benefits of taking glutathione with antibiotics as an adjunct treatment for tuberculosis infection [39], which is more common in older people. However, there is no solid evidence that glutathione supplementation alone will improve the immune function of older people.

Carnosine is an amino acid found abundantly in the skeletal muscles and brain and is synthesised from β-alanine and L-histidine [40]. Carnosine levels decrease with age and with gender, and males reportedly have higher levels stored in their skeletal muscles [41]. In our study, carnosine levels were lower in FR, HO and RA neutrophils compared to HY. Carnosine has antioxidant effects which has previously been reviewed [42]. Although there are no studies on carnosine levels in human neutrophils in ageing or frailty, there are studies on animal models or other phagocytes such as macrophages. An *in vitro* study looking at PMA-induced ROS in mouse-derived macrophages showed that carnosine decreased the intracellular concentration of superoxide anions (O_2_^−^•) as well as the expression of the NOX2 genes [43]. Supplementation of carnosine in aged rats caused a decrease in superoxide anion production and enhanced chemotactic and phagocytic activity to comparable control levels in neutrophils [44]. Carnosine suppressed LPS-induced ROS and MPO production in the lungs of mice in a model of lung injury [45]. It is widely known that cancer incidence increases with age, and in regard to tumour growth, carnosine has extensively been studied for its role in tumour suppression [46]. In cultured fibroblasts, the addition of carnosine slowed the telomere shortening rate as well as decreased the accumulation of DNA-damage compared to fibroblasts cultured without carnosine [47]. Carnosine has also been demonstrated to improve memory and inflammatory markers which are both associated with extreme old age. A 3-month interventional study of anserine and carnosine supplementation showed improved delayed recall verbal memory and a decrease in IL-8 levels in serum compared to the placebo-controlled group [48]. Additionally, in a randomised, placebo-controlled trial, the precursor of carnosine, β-alanine increased muscle carnosine levels and improved physical performance [49] as well as muscle recovery in older people (60+ years old) [50]. This could present a possible intervention for the treatment of sarcopenia prevention or attenuation.

Several metabolites regulating immune function were altered in our study. The essential amino acids lysine, valine, leucine, and isoleucine were all significantly lower in FR neutrophils compared to HY, and valine was significantly lower in HO compared to HY. Essential amino acids cannot be produced in humans and must be obtained through the diet. Neutrophils require amino acids as a fuel source and to produce and maintain intracellular proteins. A deficiency of dietary protein is common among the older population. A study utilising data from 11,680 adults from 2005-2014 showed that 46-50% of people over 60 do not meet the daily protein intake recommendation [51]. Valine, leucine, and isoleucine are branched-chain amino acids (BCAAs). A lack of BCAAs in the diet impairs aspects of the immune response and increases susceptibility to pathogens [52]. An interventional study showed that a daily dose of 12g BCAAs via oral supplementation improved phagocytic function of neutrophils and natural killer cell activity in cirrhotic patients [53]. Leucine was administered at a dose of 40mg/mL to mice infected with herpes simplex virus type 1 (HSV-1) and resulted in an increased number of IFN-γ and TNF-α-producing CD4+ T cells specific to the HSV-1 antigen [54]. Additionally, this was associated with better survival of the mice. A decrease in availability of amino acids within neutrophils could be one way in which their function declines with age. Although not significant in this study, the majority of neutrophil amino acid studies have investigated the role of glutamine due to its critical role in neutrophil metabolism. Evidence for other amino acid levels and their roles in neutrophils is extremely limited. There have been reports that neutrophils release the positively charged BCAAs valine, isoleucine, leucine, the aromatic amino acids tyrosine, phenylalanine, and the free amino acids arginine, ornithine, lysine, hydroxylysine, and histidine upon adherence to fibronectin [55]. Neutrophils bind to fibronectin in inflammatory contexts and may additionally release proteases to degrade fibronectin, clearing a path for their migration towards inflammatory sites [56]. Frailty has been described as a condition of accelerated ageing [57]. Therefore, it is reasonable to assume that the hallmarks of ageing, including cellular senescence, are also accelerated, and therefore, increased in frailty compared to healthy ageing. Neutrophils can migrate towards the SASP produced by senescent cells. In 2-year-old mice (elderly aged mice), senescent cells recruited neutrophils toward the liver via the SASP [58].

Frail neutrophils also had significantly higher GTP levels compared to the HY group. GTPases are enzymes that act as molecular switches for various cellular process such as cell migration and actin cytoskeleton remodelling. GTPases either bind to GTP or GDP based on their activation state. Activated GTPases are GTP-bound and are hydrolysed to GDP-bound enzymes to become inactivated. The GDP metabolite is not available in Chenomx, and it would have been interesting to observe the ratio of GDP to GTP in these neutrophils. Currently we do not know if we have GDP in our data, and it would be interesting future work to determine its presence and abundance. 3-Hydroxyisovalerate was also elevated in FR neutrophils compared to HO. It is a metabolite of leucine and is currently used as a biomarker for biotin deficiency [59]. Currently, there are only a handful of studies detailing the role of increased/decreased 3-hydroxyisovalerate in neutrophils, however, it has previously been reported as reduced in neutrophils from obese individuals and increased in the urine of people with neutropenia [60] [61].

As part of this study we also performed NMR lipidomics for the first time in human neutrophils, and identified significant changes in neutrophil lipids with frailty status. Our bespoke pattern file was annotated based on matching to literature for specific lipid classes as there is currently no definitive NMR lipid library available. Although there are a few studies identifying neutrophil lipid metabolites using ^1^H-NMR spectroscopy [62] [63], this is the first study that seperates the polar and lipid fractions of neutrophils in this particular way (reserving the lipid pellet after polar extraction and then dissolving directly in chloroform). This protocol was adapted from a previous study that utilised ^1^H-NMR spectroscopy at 700MHz to profile lipids in zebrafish embryos [12]. The significant lipid bins we identified correlated to total cholesterol and the omega 6 fatty acids (which includes arachidonic acid (AA)). Total cholesterol and AA were only significantly different between FR and HY suggesting a change with age rather than frailty status. Notably, FR neutrophils had significantly higher abundance of total cholesterol (free cholesterol and cholesterol esters). It has been known for decades that neutrophils have cholesterol receptors, as well as the ability to uptake cholesterol [64]. Previous studies have reported that membrane cholesterol levels within neutrophils are linked to their function [65]. Lipid rafts, also known as microdomains, are proteins and lipid structures enriched in cholesterol and sphingolipids that float within the bilayer of cell membranes [66]. They serve as environments upon which receptors are rapidly recruited to the plasma membrane in cell activation. One example is TLR4 which is recruited to lipid rafts upon ligand biding of stimuli such as LPS. Lipid raft distribution is disorganised with ageing, consequently disrupting downstream signalling events in neutrophils from older people [67]. We also identified lower choline levels in FR neutrophils. The exact function of choline within neutrophils is not known, however, it is known that choline is the precursor for acetylcholine. When activated, neutrophils produce acetylcholine which can feedback to neuronal networks and also signal to other immune cells [68]. Although neutrophils can produce acetylcholine, they synthetise lower amounts of acetylcholine compared to other lymphocytes [69]. In bovine neutrophils, phagocytic and killing capacity of neutrophils *in vitro* were diminished with increasing doses of choline, whereas lymphocyte proliferation was linearly enhanced [70]. In a mouse model of hyperalgesia (increased sensitivity to pain), injections of choline prevented inflammatory pain nociception without affecting neutrophil migration or cytokine production [71]. Choline is also linked to lipid homeostasis, and in particular lower levels of circulating choline in mice can lead to accumulation of cholesterol in the liver [72] and higher circulating levels of HDL and non-HDL cholesterol [73] suggesting a possible link between choline and cholesterol levels in neutrophils from people with frailty.

Due to ethical restrictions the blood samples collected in this study were not fasted. It was also not possible to record the use of statins for this study so the differences in cholesterol levels could be due to differences in diet or medication usage, and it would be beneficial to include recording of statin treatments in future studies. It would also be interesting in future to see if there would be an opportunity to collect fasted blood and whether this would yield the same results. Neutrophils are capable of synthesising, and esterifying cholesterol. Cholesterol sulfate (CS) is a derivative of cholesterol that is produced in neutrophils as well as many other cells and tissues [74]. One study showed that CS directly binds to the catalytic DHR-2 domain of DOCK2, inhibiting its Rac-GEF activity, and suppressing neutrophil and T-cell migration [75]. It is known that certain tissues limit or suppress immune cell infiltration, such as the eyes. In mice, it was found that CS was produced by the gland that secretes the lipids that form the outer layer of the tear film covering the eye [75]. Another study observed the inhibition of leukotriene synthesis by cholesterol esters, CS and cholesterol phosphate [76]. Both CS and cholesterol also inhibited AA release from neutrophils [76]. According to the current literature, cholesterol and its derivatives can have an inhibitory effect on neutrophil migration. This could be one way in which cell migration becomes dysregulated in older neutrophils [77].

Omega 6 fatty acids (which include AA), were higher in HY neutrophils compared to FR. Unlike omega 3 fatty acids which are considered to be anti-inflammatory, omega 6 fatty acids are considered pro-inflammatory [78]. This difference between the HY and FR neutrophils could be due to differences in diet. Omega 3 and omega 6 fatty acids are stored in the cell membrane of neutrophils. Previous randomised placebo-controlled trials have shown that supplementation of omega 3 increased the percentage of omega 3s and reduced the percentage of AA in the membrane of in human neutrophils [79] [80]. AA is the precursor for eicosanoids, which are lipid mediators and includes leukotrienes, lipoxins, and prostanoids among others [81]. Eicosanoids are synthesised from the oxidation of AA via the cyclooxygenase (COX), lipoxygenase (LOX), and cytochrome P450 enzymes. Eicosanoids are generally associated with inflammation and AA has previously earned the title of the most crucial precursor of inflammatory pathways [82]. Anti-inflammatory medications such as corticosteroids and non-steroidal anti-inflammatory drugs (NSAIDs) target the COX and LOX enzymes to reduce inflammation as well as pain [83]. The COX-2 enzyme expression is also increased in tumours [84] and COX/LOX inhibitors have been proposed as promising treatments for various cancers [85]. The other lipid mediators produced by the AA pathways are leukotrienes and lipoxins. Leukotriene B4 (LTB4) is one of the strongest lipid chemoattractant compounds produced in neutrophils and macrophages [86]. These results could suggest that the HY neutrophils have a greater capacity to induce chemotaxis due to increased intracellular LTB4.

The more likely explanation for the increased level of omega 6 fatty acids in HY neutrophils is the production of lipoxins which are anti-inflammatory mediators. Lipoxin A4 and lipoxin B4 treatment in whole blood from atherosclerosis patients attenuated neutrophil oxidative burst, a key contributor to atherosclerotic development [87]. Lipoxin A4 also reduced ROS production through the inhibition of NOX enzymes in microglial cells [88]. In a rat model of diabetes, lipoxin A4 inhibited the expression levels of inflammatory cytokines including TNF-α, IL-1β, and IL-18 [89]. The administration of lipoxin A4 in a rat model of acute respiratory distress syndrome reduced the degree of lung injury and promoted the phagocytosis and apoptosis of neutrophils [90]. There were no effects observed in a control model of rats with ARDS and neutropenia. *In vitro*, lipoxin A4 has been identified as a stop signal for neutrophil swarming, whereas LTB4 aids swarming [91]. Dysregulation in either of these lipids could greatly affect the outcome of the immune response. The ratio of metabolites derived from omega 6 fatty acids including AA would allow us to gain valuable information on the pathway differences between healthy younger people and people with frailty that we currently cannot define using NMR spectroscopy alone. The neutrophils analysed in our study were isolated from people without infections. Investigating the differences between neutrophil lipid mediators between people of different ages with and without infections could provide an insight into whether a lipid-mediator imbalance exists in aged people. Stimulating neutrophils with pro-inflammatory compounds from cohorts of different ages could also provide an insight into which pathways and mediators are increased in ageing compared to younger people. Additionally, there is a possibility that these differences are affected by the diet, fasting blood could be used in future studies to exclude any changes observed through diet.

As part of this study we wanted to test our hypothesis that frailty is an inflammatory condition, and we did this by including samples from people with RA in our study design. Interestingly, there were only two significant metabolite differences between FR and RA, NADP and isoleucine. NADP also distinguished FR from the HY and HO groups. Metabolites that were significantly different between HO and HY include the antioxidants glutathione and carnosine, however, there were no differences for these metabolites between FR and RA. This suggests that RA neutrophils, similarly to HO and FR, have a reduced antioxidant capacity compared to HY neutrophils. Interestingly, the metabolites elevated but not statistically significant (VIP>1 model 60.4% accuracy) in FR compared to RA included ATP, ADP, AMP, and taurine. Neutrophils naturally have high levels of taurine, and these are even higher in RA neutrophils compared to healthy controls [9]. The taurine levels of neutrophils and monocytes are sharply decreased after LPS exposure, and taurine supplementation was found to repress NET formation in a ROS-dependent pathway in neutrophils [92]. ATP is hydrolysed into ADP or further to AMP for energy release. ATP levels drop significantly from 1.9 to 1.0fmol/cell during neutrophil phagocytosis, whereas the rate of glycolysis remains unchanged [93]. These studies suggest that RA neutrophils could be more metabolically active compared to FR neutrophils since they have a decreased level of these metabolites. This would make sense as RA is considered a pro-inflammatory condition, however, it does not mean that FR is not inflammatory, rather it is less inflammatory than the RA disease.

One limitation of the study design was that whilst the classification criteria for frailty includes age >65 years, the classification age range for the RA group was anyone above 18 years old with diagnosed RA. For future work, it would make a more appropriate comparison to include only age-matched people with RA as a positive control group. The changes seen with age could be skewing the current comparison between RA and FR. For example, isoleucine was higher in RA and HY, and this difference could be from the younger participants in the RA group. With a more distinct age group for the RA group, any changes observed could be solely due to the different disease states without the influence of age. To mitigate this the RA group was removed from our ANOVA and this identified additional significant metabolite differences, indicating the need for high powered studies (more samples less groups) and/or age-matching of FR and RA participants in the future [94]. Other limitations in the study included the use of non-fasted study participants. For example, the difference observed in the essential amino acids leucine, isoleucine, lysine, and valine may be due to diet and/or fasting status. The BMI, smoking status, exercise levels and medication use was not recorded for this study. We also did not record the use of statins, or food supplements such as omega-3 fish oils, as mentioned earlier. These all have the potential to influence metabolism within neutrophils since they each have the potential to affect immune function to certain degrees. For future studies, it would be interesting to see whether the results are the same when blood samples are fasted overnight prior to neutrophil isolation. Utilising other analytical platforms such as liquid chromatography or gas chromatography coupled mass spectrometry, which have a broader range of metabolite detection, could also aid with identifying more metabolites in order to get the most out of these data.

In summary, this study has identified metabolic differences in neutrophils associated with healthy ageing, and ageing with frailty. We have also identified significant overlap between fail neutrophil metabolites and those found in RA neutrophils, suggesting an inflammatory environment in frailty. It is widely accepted that, due to the complexity of the ageing process, no single biomarker could accurately measure the differences in age among humans. However, we identified intracellular neutrophil NADP as a unique biomarker of frailty. It is likely that a combination of biomarkers could be identified and used as a single parameter to distinguish differences between humans of different ages, and also, how this affects the cells from which they were extracted. In the last decade in particular, there has been enormous growth in our understanding of cell metabolism, immunometabolism and the wider role of metabolites as signalling molecules. Despite this, metabolites within neutrophils have only recently gained interest. This is likely due to their relatively short lifespan, cell culture challenges, and historic misconception of their metabolic simplicity. Inflammatory and metabolic pathways are interdependent. Further investigation into neutrophil metabolism and metabolites could provide an insight into how and why they become dysregulated with ageing and frailty, leading to reduced protection from infections in older people.

## Data Availability

All data produced in the present study are available upon reasonable request to the authors

## Acknowledgements

We would like to thank the clinical staff at Liverpool University Hospital NHS Foundation Trust for help recruiting people with frailty and RA for this study. We also thank the Liverpool Shared Research Facilities (LivSRF) for support and training in statistical analysis provided by the high-field NMR facility and Computational Biology Facility (CBF) by Dr Rudi Grosman and Dr Eva Caamano Gutierrez.

## Authorship Declaration

G.A.A. performed the experiments, formal analysis, methodology, visualisation, writing – original draft. G.F. methodology, writing – review and editing. L.G. methodology, writing – review and editing. A.A. supervision, participant screening and recruitment, writing – review and editing. M.M.P. conceptualisation, methodology, data curation, formal analysis, supervision, funding acquisition, visualisation, writing – original draft. H.L.W. conceptualisation, funding acquisition, project administration, supervision, visualisation, writing – original draft.

## Conflicts of interest

None

### Abbreviations

AA: arachidonic acid;
ADP: adenosine diphosphate;
ANOVA: analysis of variance;
ATP: adenosine triphosphate;
BCAA: branched-chain amino acids;
BH: Benjamini-Hochberg;
COX: cyclooxygenase;
CRS: correlation reliability score;
CS: cholesterol sulfate;
FDR: false-discovery rate;
FR: frailty;
GDP: guanosine diphosphate;
GTP: guanosine triphosphate;
HDL: high-density lipoprotein;
HO: healthy older;
HY: healthy younger;
IFN: interferon;
LOX: lipoxygenase;
LPS: lipopolysaccharide;
LTB4: leukotriene B4;
MPO: myeloperoxidase;
MSEA: metabolites set enrichment analysis;
NADP: Nicotinamide adenine dinucleotide phosphate (oxidised form);
NADPH: Nicotinamide adenine dinucleotide phosphate (reduced form);
NET: neutrophil extracellular trap;
NMR: nuclear magnetic resonance;
NSAID: non-steroidal anti-inflammatory;
OXPHOS: oxidative phosphorylation;
PLS-DA: partial least squares-discriminant analysis;
PQN: probabilistic quotient normalization;
RA: rheumatoid arthritis;
ROS: reactive oxygen species;
SASP: senescence-associated secretory phenotype;
SMPDB: small molecule pathway database;
TCR: T-cell receptor;
TLR: Toll-like receptor;
VIP: variable importance of projection

