## Supplementary Data for "1H NMR metabolomics and lipidomics analysis of neutrophils reveals biomarkers of ageing, inflammageing and frailty"

**^1^Institute of Life Course and Medical Sciences, University of Liverpool, 6 West Derby Street, Liverpool L7 8TX, UK**

^2^Bunbury Regional Hospital, Bunbury 6230, Western Australia

^3^Division of Internal Medicine, University of Western Australia, Crawley 6009, Western Australia

**^4^High-Field NMR Facility, Liverpool Shared Research Facilities (LivSRF) University of Liverpool, Crown Steet, Liverpool L69 7ZB, UK**

**^5^Institute of Systems, Molecular and Integrative Biology, University of Liverpool, Crown Street, Liverpool L69 7ZB, UK**

| **Supplementary Table 1.** Participant demographics for each cohort of neutrophil samples | | | | |
| --- | --- | --- | --- | --- |
|  | **Frail (FR)** | **Healthy Older (HO)** | **Healthy Younger (HY)** | **Rheumatoid Arthritis (RA)** |
| **Sample size** | 31 | 24 | 21 | 16 |
| **Male (%)** | 39 | 38 | 38 | 19 |
| **Female (%)** | 61 | 62 | 62 | 81 |
| **M:F ratio** | *p*>0.05 | *p*>0.05 | *p*>0.05 | *p<*0.05 |
| **Average age** | 84 | 66 | 22 | 55 |
| **Age matching pval** | FR:HO  *p<*0.05 | FR:HY  *p<*0.05 | HY:HO  *p<*0.05 | RA:FR  *p<*0.05 |

**Supplementary Table 2**. **Representative metabolite peaks.** Scores are rounded to the nearest percent. Scores with “na” represent metabolites with a single peak annotated and therefore, that single peak was chosen as the representative. * = Metabolite has more than one expected peak, however, due to overlap with higher concentration metabolites or low signal, these peaks were not annotated (less than 20% total peak area – see **figure 3.2** for further details). 1** = level 1 confirmation via 2D spectra. *** Denotes representative peaks that are overlapping with at least one more metabolite. All metabolites annotated were naturally occurring isomers; amino acids these are L-isomers, for saccharides such as glucose these are D-isomers.

| HMDB ID | Metabolite Name | Representative Peak (ppm) | Correlation Score (%) | MSI Level of Identification |
| --- | --- | --- | --- | --- |
| HMDB0000011 | 3-Hydroxybutyrate | 2.4227…2.38831*** | na | 2 |
| HMDB0000754 | 3-Hydroxyisovalerate | 1.2578…1.2481 | na | 2 |
| HMDB0031645 | Acetamide | 1.9769…1.9726 | na | 2 |
| HMDB0031523 | Acetate | 1.927…1.9106 | na | 2 |
| HMDB0304256 | Acetoacetate | 2.26762…2.26154 | na | 2 |
| HMDB0001659 | Acetone | 2.239…2.23255 | na | 2 |
| HMDB0001341 | ADP | 8.5552…8.5465 | 92 | 2 |
| HMDB0000161 | Alanine | 1.4931…1.4723 | 90 | 1** |
| HMDB0000045 | AMP | 8.62035…8.61231 | 76 | 2 |
| HMDB0000517 | Arginine | 1.7559…1.5705 | na* | 2 |
| HMDB0000168 | Asparagine | 2.88274…2.87228 | 90 | 2 |
| HMDB0000191 | Aspartate | 2.83492…2.8235 | 99 | 2 |
| HMDB0000538 | ATP | 8.2809…8.2608 | 99 | 2 |
| HMDB0304270 | Benzoate | 7.52953…7.46828 | 98 | 2 |
| HMDB0000033 | Carnosine | 3.073…3.04119 | 99 | 2 |
| HMDB0000097 | Choline | 3.2123…3.20402 | na | 2 |
| HMDB0003417 | Cysteine | 4.03953…4.023 | 100 | 2 |
| HMDB0000087 | Dimethylamine | 2.73513…2.73007 | na | 2 |
| HMDB0000142 | Formate | 8.46372…8.45373 | na | 2 |
| HMDB0000122 | Glucose | 3.24254…3.23577 | 73 | 2 |
| HMDB0000148 | Glutamate | 2.1539…2.12588 | 86 | 2 |
| HMDB0000641 | Glutamine | 2.49037…2.4227 | 92 | 1** |
| HMDB0000125 | Glutathione | 2.993…2.9605 | 92 | 1** |
| HMDB0000131 | Glycerol | 3.57906…3.57116 | 98 | 2 |
| HMDB0001273 | GTP | 8.14861…8.14039 | 95 | 2 |
| HMDB0000177 | Histidine | 7.83434…7.8277 | na | 2 |
| HMDB0000719 | Homoserine | 2.01057…2.00828 | 87 | 2 |
| HMDB0000175 | IMP | 8.21929…8.21087 | 76 | 2 |
| HMDB0000671 | Indolelactate | 7.77012…7.73072 | na | 2 |
| HMDB0001873 | Isobutyrate | 1.0731…1.0564 | Na | 2 |
| HMDB0000172 | Isoleucine | 1.02566…1.00515 | 95 | 2 |
| HMDB0000863 | Isopropanol | 1.1841…1.1699 | na | 2 |
| HMDB0000190 | Lactate | 4.1316…4.0958 | 96 | 1** |
| HMDB0000687 | Leucine | 0.9809…0.9521 | 91 | 2 |
| HMDB0000182 | Lysine | 3.04119…2.993 | 96 | 2 |
| HMDB0000696 | Methionine | 2.66588…2.65582 | 99 | 2 |
| HMDB0000211 | Myoinositol | 3.6413…3.61922 | 88 | 2 |
| HMDB0000902 | NAD | 8.18161…8.17652 | 64 | 2 |
| HMDB0000217 | NADP | 8.20118…8.19462*** | na* | 2 |
| HMDB0000159 | Phenylalanine | 7.4459…7.415 | 83 | 2 |
| HMDB0001565 | Phosphocholine | 3.21757…3.21274 | na | 2 |
| HMDB0000162 | Proline | 4.08688…4.0759 | 78 | 2 |
| HMDB0000267 | Pyroglutamate | 2.5174…2.49113 | 80 | 2 |
| HMDB0000279 | Saccharopine | 2.12588…2.08299 | 71 | 2 |
| HMDB0000271 | Sarcosine | 2.7631…2.75409 | 100 | 2 |
| HMDB0000187 | Serine | 3.96478…3.95065 | 96 | 2 |
| HMDB0000251 | Taurine | 3.28559…3.26545 | 100 | 2 |
| HMDB0000167 | Threonine | 3.60498…3.5792 | na | 2 |
| HMDB0000158 | Tyrosine | 6.91501…6.89646 | 98 | 2 |
| HMDB0000883 | Valine | 1.0046…0.9881 | 98 | 2 |

**Supplementary Table 3. Enriched metabolic pathways for neutrophil polars.**

| **Pathway** | **Total number of metabolites in pathway** | **Metabolites matched** | **FDR** |
| --- | --- | --- | --- |
| **FR - HO** | | | |
| Aspartate metabolism | 35 | 5: acetate, asparagine, aspartate, GTP, indolelactate | 0.0126 |
| Ketone body metabolism | 13 | 3: 3Hydroxybutyrate, acetate, NAD | 0.0345 |
| Ammonia recycling | 31 | 4: asparagine, aspartate, histidine, NAD | 0.0345 |
| Beta-alanine metabolism | 34 | 4: histidine, aspartate, NADP, NAD | 0.037 |
| **FR - HY** | | | |
| Glutathione metabolism | 20 | 4: glutathione, alanine, NADP, cysteine | 0.029 |
| **HO - HY** | | | |
| Glycine and serine metabolism | 59 | 6: serine, sarcosine, arginine, cysteine, methionine, NAD | 0.0105 |
| Methionine metabolism | 42 | 5: serine, sarcosine, cysteine, methionine, NAD | 0.011 |
| Ammonia recycling | 31 | 4: asparagine, serine, aspartate, NAD | 0.0264 |
| **FR - RA** | | | |
| Pyruvate metabolism | 47 | 6: AMP, glutathione, NADP, ATP, GTP, ADP | 0.00425 |
| Valine, Leucine and Isoleucine degradation | 59 | 6: acetate, isoleucine, valine, leucine, ADP, ATP | 0.00591 |
| Glutathione metabolism | 20 | 4: glutathione, NADP, ADP, ATP | 0.00591 |
| Thiamine metabolism | 9 | 3: ATP, ADP, AMP | 0.0065 |
| Glycerolipid metabolism | 25 | 4: glycerol, NADP, ADP, ATP | 0.00882 |
| Glutamate metabolism | 48 | 5: ATP, AMP, ADP, NADP, glutathione | 0.00955 |
| Urea cycle | 28 | 4: AMP, ATP, ADP, arginine | 0.00955 |
| Pentose phosphate pathway | 29 | 4: AMP, ADP, ATP, NADP | 0.00955 |
| Arginine and Proline metabolism | 52 | 5: AMP, NADP, ADP, ATP, arginine | 0.00955 |
| Fructose and mannose degradation | 31 | 4: NADP, ATP, ADP, GTP | 0.0103 |

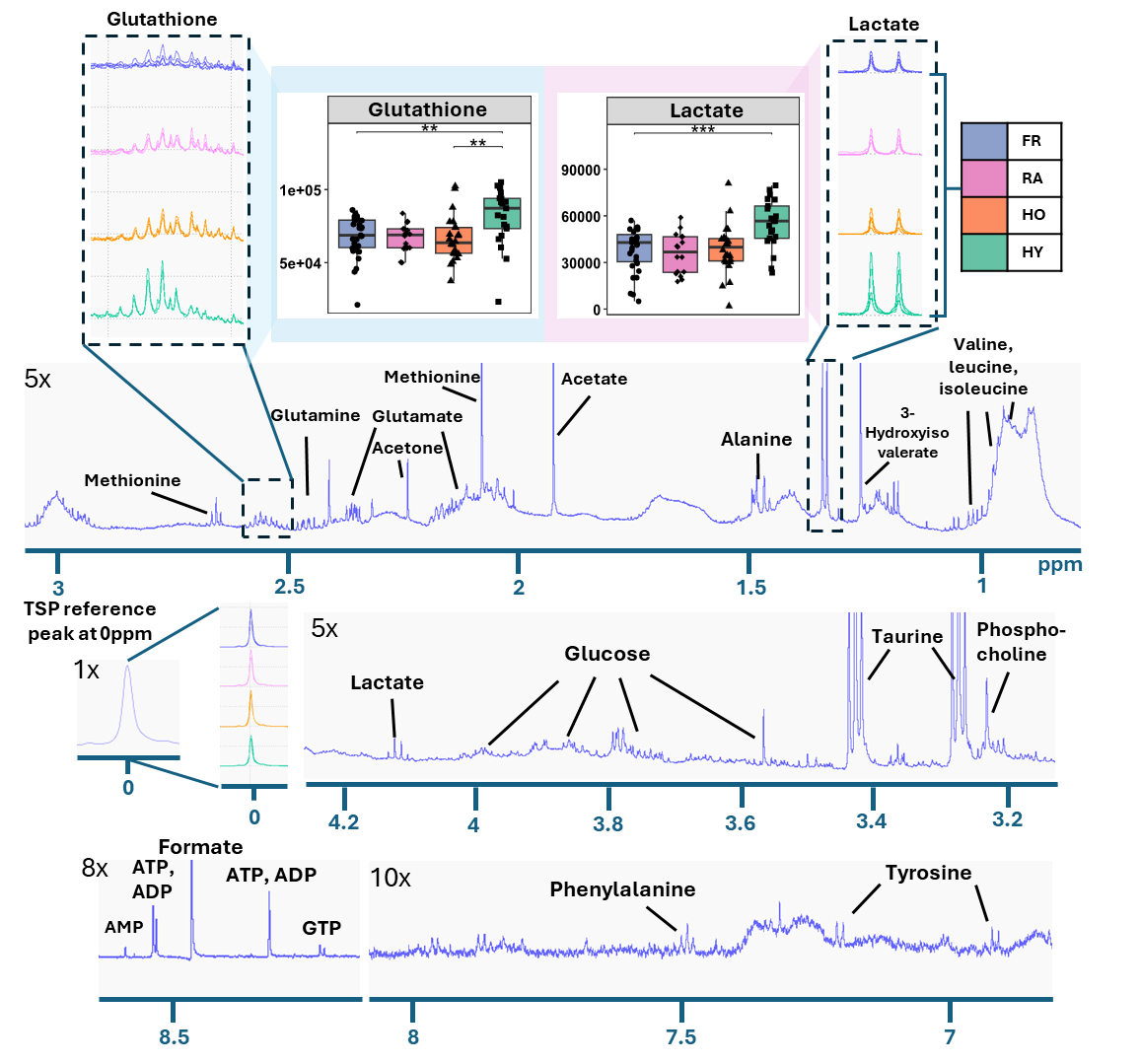

**Supplementary Figure 1.** **700MHz ^1^H-NMR spectra annotation of polar neutrophil metabolites on a representative healthy young control spectrum.** Differences were seen for some metabolites in the raw spectra before processing, for example glutathione and lactate which were significantly different after analysis via ANOVA.

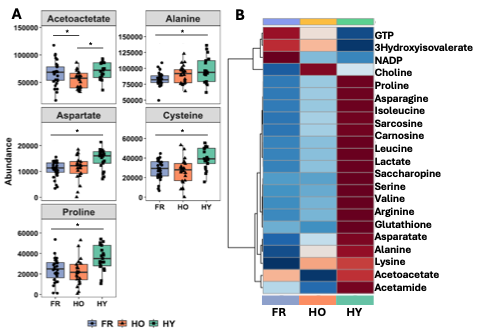

**Supplementary Figure 2. One-way ANOVA results of metabolic data between FR and the healthy groups.** Excluding RA resulted in an additional 5 significantly different metabolites shown by ANOVA, shown in (A) boxplots for alanine, acetoacetate, proline, cysteine, and aspartate (*p<0.05). (B) Heatmap of significant metabolites between FR, HO, and HY only. Metabolites were log10 transformed and displayed as mean intensities of blue or red. Blue shows lower than mean abundance and red shows higher than mean abundance.

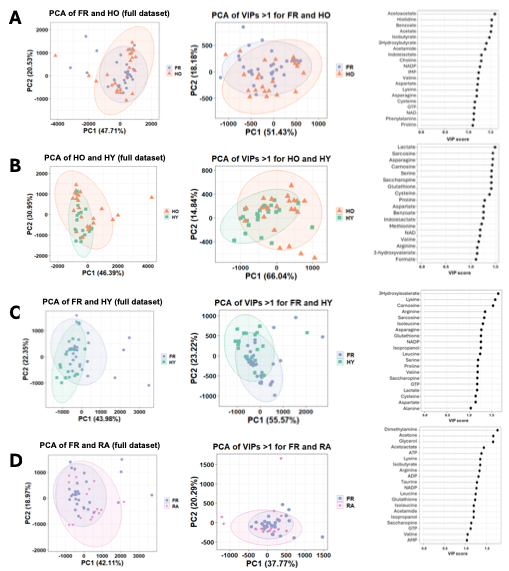

**Supplementary Figure 3. Analysis of metabolites influential to PLS-DA models.** PCA scores plots for full dataset (left) and subset for influential metabolites (VIP>1, middle) along with VIP scores plot (right). (A) FR vs HO, (B) HO vs HY, (C) FR vs HY, and (D) FR vs RA. Neutrophils were isolated from participants who were healthy old (HO, orange triangles, n=24), frail (FR, blue circles, n=31), healthy young (HY, green squares, n=21) or rheumatoid arthritis (RA, pink, n=16).
